# A pilot study of the mutational landscape of colorectal cancer in Black South Africans

**DOI:** 10.64898/2026.11.18.26368225

**Authors:** Brandenburg Jean-Tristan, Campbell Lisa, Engelbrecht Dewaldt, Mathew G. Chistopher, Gaynor Thompson, Darisia Moonsamy, Nadine Harran, Martin Richard M, Bebington Brendan, Soldati Vuyo, Perdomo Sandra, Stevens Wendy, Fabian June, Haycock Philip C, the MOSAIC collaboration

## Abstract

**Background:** Colorectal cancer (CRC) is a public health concern in sub-Saharan Africa, where the molecular landscape of the disease remains poorly characterised. In particular, genomic studies of resident Black African populations are limited, and the contribution of population-specific mutational processes and potential infectious exposures remains insufficiently understood. The Molecular, Somatic and Infectious Drivers of Colorectal Cancer in Africa (MOSAIC) collaboration aims to address these gaps by generating population-specific genomic data. This pilot study aimed to assess the feasibility of paired tumour-normal whole-genome sequencing (WGS) and to provide an initial characterisation of somatic mutations and mutational signatures in South African patients with CRC.

**Methods:** We recruited five Black South African patients with newly diagnosed, treatment-naïve CRC through the CoRecT Cancer study at Chris Hani Baragwanath Academic Hospital And Wits University Donald Gordon Medical Centre, Johannesburg. Fresh-frozen colorectal tumour tissue and matched peripheral blood samples were subjected to paired-end WGS (2 × 151 bp) using the Illumina NovaSeq X Plus platform. Somatic variants were identified using CaVEMan and Strelka2, followed by functional annotation using the Ensembl Variant Effect Predictor (VEP). Protein-altering variants were characterised using the R package maftools. Mutational signature contributions were estimated using SigProfilerAssignment and the COSMIC v3 reference catalogue.

**Results:** All ten sequencing samples, comprising five tumour-blood pairs, passed quality control. Estimated sequencing depth ranged from 49.2X to 82.4X for tumour samples and from 22.7X to 40.7X for matched blood samples, with a median of 96.3% well-aligned reads. Following somatic variant calling and filtering, the number of retained mutations ranged from 1,106 to 65,379 per tumour. Three of the five tumours harboured at least one predicted protein-altering variant, comprising seven missense and four splice-site variants across eleven genes. Mutational signature analysis of single base substitutions (SBS) revealed inter-individual heterogeneity. SBS1 and SBS5 were detected across all five tumours, while additional signatures included SBS18, SBS20, SBS26 and SBS44, among others. Several tumours exhibited signatures associated with mismatch repair-related mutational processes.

**Conclusions:** This pilot study demonstrates the feasibility of paired tumour-normal WGS and preliminary somatic mutation profiling in Black South African patients with CRC. The findings provide initial insights into the heterogeneous mutational landscape of CRC in this underrepresented population. However, the small sample size and incomplete clinical and molecular characterisation necessitate cautious interpretation. Larger studies incorporating comprehensive mismatch repair assessment, copy-number and structural variant analysis, germline susceptibility profiling and investigation of infectious exposures will be essential to validate these preliminary observations and better understand the molecular and environmental drivers of CRC in African populations.

## Background

Colorectal cancer (CRC) is the third most common cancer in men and women in South Africa and the fifth most common cancer in sub-Saharan Africa (Amer et al., 2024). Notably, CRC is diagnosed at a younger age in Black African populations compared with European descent populations (Alatise et al., 2021), with a study from South Africa reporting differences of approximately 10 years in median age at diagnosis between European descent and African descent in South Africa (Motsuku et al., 2021) . Despite this growing burden, the molecular landscape of CRC in sub-Saharan African populations remains poorly characterised. Among the studies conducted in the region are targeted sequencing investigations in Nigerian patients (Alatise et al., 2021) and whole-exome sequencing studies in South African patients (Aldera et al., 2025).

Sub-Saharan Africa experiences a substantial burden of infectious diseases, including HIV, which may influence host immunity, inflammation, and the composition of the gut microbiome. South Africa, in particular, has one of the highest HIV burdens globally (HIV/AIDS, 2026) . Although the contribution of HIV to colorectal carcinogenesis remains uncertain, bacterial components of the gut microbiome have emerged as potential contributors to tumour initiation and progression (J. Kim & Lee, 2022).

Evidence implicates colibactin-producing polyketide synthase genomic island (pk+) *Escherichia coli* in colorectal carcinogenesis. Colibactin is a bacterial genotoxin capable of inducing DNA damage, DNA crosslinks, and genomic instability (Dziubańska-Kusibab et al., 2020; Pleguezuelos-Manzano et al., 2020). Experimental exposure of human intestinal organoids to pks+ *E. coli* has demonstrated a characteristic mutational signature that has been identified in human colorectal tumours. Subsequent studies have characterised associated mutational signatures, including SBS88 and ID18. Importantly, an international whole-genome sequencing study identified geographical variation in mutational processes and enrichment of colibactin-associated CRC (Díaz-Gay et al., 2025) .

Somatic mutations play a central role in colorectal carcinogenesis through the accumulation of alterations affecting oncogenes, tumour suppressor genes and other cancer-related pathways. Beyond individual mutations, mutational signatures provide insights into the biological processes and environmental exposures contributing to cancer development. Characterising these signatures in African populations may therefore reveal mutational processes that remain underrepresented in international cancer genomic studies.

This study emerged from a partnership between the University of the Witwatersrand (Wits), Johannesburg, and the University of Bristol. Following a two-day cancer workshop held in Johannesburg, South African researchers and stakeholders, together with UK and other international partners, including representatives from the International Agency for Research on Cancer (IARC), the Global Colon Cancer Association (GCCA) and the University of Botswana, came together to discuss research priorities and unmet needs in CRC research and detection in southern Africa.

The discussions highlighted several important gaps in CRC research across South Africa and the wider African continent, particularly the limited understanding of the genetic landscape, molecular pathways and potential infectious drivers of CRC in Black African populations. We founded the Molecular, Somatic and Infectious Drivers of Colorectal Cancer in Africa (MOSAIC) collaboration to address these gaps. One of the main aims of MOSAIC is to strengthen research capacity and generate population-specific genomic data to better understand CRC aetiology and identify potential opportunities for improved detection and prevention.

As an initial proof-of-concept study within the MOSAIC collaboration, we performed paired sequencing of colorectal tumour tissue and matched peripheral blood from five patients. This pilot aims to establish the feasibility of identifying somatic mutations, characterising affected cancer-related genes, and exploring mutational signatures, including those potentially associated with bacterial exposures such as colibactin. These preliminary findings will provide a foundation for larger-scale investigations into the molecular, somatic and infectious drivers of CRC in African populations.

## Methods

### Samples and collections

Participants were recruited in July 2026 through the CoRecT Cancer study, an ongoing hospital-based colorectal cancer recruitment initiative at Chris Hani Baragwanath Academic Hospital (CHBAH) and Wits University Donald Gordon Medical Centre (WDGMC) in Johannesburg, South Africa. CoRecT Cancer is a biorepository study aimed at building a resource for CRC research in South Africa. Eligible participants were adults (aged 18 years or older) of Black South African ancestry with newly diagnosed primary colorectal adenocarcinoma who had not received previous cancer treatment. Patients with a history of chemotherapy, radiotherapy or surgery prior to enrolment were excluded. Potential participants were identified through routine clinical investigations and referred by their treating clinicians. Eligibility was assessed by the study team before enrolment and obtaining informed consent. For this pilot study, five patients meeting these criteria were selected for paired tumour (Biopsy at the time of first diagnosis or collected at the time of surgical resection) and peripheral blood analysis. Tumour tissue was collected during routine biopsy and colonoscopy procedures and from resection specimens, while peripheral blood was collected. Blood samples served as matched constitutional DNA references to distinguish somatic alterations in tumour tissue from germline variants.

For each participant, demographic, socioeconomic, familial, lifestyle, clinical and pathological information was collected using REDCap (Harris et al., 2009) and compliant to Protection of Personal Information Act (POPI Act). Demographic, socioeconomic, lifestyle and medical information were collected. For participants, pathological information included tumour location, histological type and grade, extent of tumour invasion, perineural invasion, tumour budding, lymph node involvement, tumour deposits and surgical margin status. Clinical staging information was recorded using radiological TNM classifications (Amin et al., 2017). Information on mismatch repair protein expression, microsatellite instability and BRAF V600E testing was also collected on the histology specimens where available

### Whole-genome sequencing

Genomic DNA was extracted from five fresh-frozen colorectal tumour tissue specimens using the NucleoSpin Tissue Mini Kit (Macherey-Nagel), according to the manufacturer’s instructions, with an approximate elution volume of 100 µL. Matched peripheral blood-derived DNA samples had been extracted using the Chemagic 360 automated nucleic acid extraction platform in combination with the manufacturer-specific Chemagic Buffy Coat Kit. This extraction method is based on magnetic bead technology, whereby genomic DNA is selectively bound to magnetic beads, enabling its separation and purification from other cellular components and potential contaminants. DNA concentration was quantified using the Qubit dsDNA High Sensitivity assay, while DNA purity was assessed using NanoDrop spectrophotometry. Sequencing libraries were prepared from tumour and matched normal DNA using the Illumina DNA PCR-Free Library Prep Tagmentation Kit with unique dual indexes, according to the manufacturer’s protocol. Paired-end whole-genome sequencing (WGS; 2 × 151 bp) was performed on an Illumina NovaSeq X Plus platform using a 5B flow cell at the Wits Diagnostic Innovation Hub (Wits DIH), University of the Witwatersrand, Johannesburg, South Africa. The target sequencing depths were approximately 50× for tumour samples and 25× for matched normal samples.

### Alignment, Quality control and calling

Sequencing quality assessment was done using FastQC software (v0.11.3) to assess the quality of raw sequencing reads (Andrews, 2010) Paired-end tumour and matched normal DNA sequencing reads were aligned to the human reference genome GRCh38 using NVIDIA Parabricks (v4.6.0), executed through Apptainer. Alignment was performed using the fq2bam workflow, with one GPU and five CPU threads. GPU-accelerated sorting and BAM writing were enabled using the --gpusort and --gpuwrite options, respectively. Temporary files were stored in the designated temporary directory. (Zhu et al., 2025). Statistics of alignment were estimated using samtools stats (Li et al., 2009).

Somatic variants were identified using two complementary variant callers, CaVEMan and strelka2 (S. Kim et al., 2018). CaVEMan was executed using the cgpCaVEManWrapper (v1.18.3) (Jones et al., 2016). Common variants were identified and retained as described in (Díaz-Gay et al., 2025). The analysis incorporated GRCh38-specific CaVEMan flagging and annotation resources, including VAGrENT annotation resources. Default tumour and normal copy-number values were set to 4 and 2, respectively, using the --tum-cn-default and --norm-cn-default parameters. Sample-specific copy-number and ignore-region files, together with a predefined flagging configuration, were supplied to the caller.

Strelka2 (Kim et al., 2018) was used for somatic variant calling in matched tumour-normal pairs. The workflow was configured using the tumour BAM, normal BAM and GRCh38 reference FASTA files. Variant calling was subsequently performed using local execution (-m local) with the specified number of computational threads (-j). Strelka2 SNV calls were subsequently filtered to retain variants passing the caller’s PASS filter using bcftools (bcftools view -f PASS). The resulting compressed VCF files were indexed using bcftools. CaVEMan PASS-filtered VCF files were also indexed for downstream analyses.

### Somatic variant annotation and functional analysis

Somatic variants were annotated using the vcf2maf Perl pipeline, incorporating Variant Effect Predictor (VEP, version 116) (McLaren et al., 2016) and the GRCh38 (hg38) human reference genome. Annotated variants were converted into MAF format and subsequently analysed using the R package maftools (Mayakonda et al., 2018). Downstream analyses were restricted to variants with potentially functional effects, including missense mutations, nonsense mutations, splice-site mutations, translation start-site mutations and nonstop mutations. Mutation profiles were summarised using plotmafSummary and the distribution of the 20 most frequently mutated genes across tumour samples was visualised using oncoplot from maftools library (Mayakonda et al., 2018).

### Mutational signature

Mutational signature analysis was performed using SigProfilerMatrixGenerator to generate single-base substitution (SBS), doublet-base substitution (DBS), and mutation catalogues (Bergstrom et al., 2019). Given the limited number of tumour samples, de novo mutational signature extraction was not performed. Instead, the contribution of previously described COSMIC v3 mutational signatures was assessed using Analyze.cosmic_fit from SigProfilerAssignment (Díaz-Gay et al., 2023) based on the SBS96 mutation classification framework. Signature contributions were estimated by fitting the observed mutational profiles to the reference COSMIC signature catalogue (Sondka et al., 2024). Individual signature assignments taking into account the number of somatic mutations per tumour and the confidence of the signature attributions. that signature within each tumour. Only signatures accounting for at least 5% of the total attributed mutations in a sample were retained for reporting.

## Results

### Cohort description

Five individuals were included in the pilot study: four females and one male (Table 1). Their ages ranged from 34 to 73 years, with a median age of 54 years. A family history of cancer was reported for one individual, while three reported no family history and information was unknown for one. Tumour location was recorded for three individuals: two had caecal tumours and one had a sigmoid colon tumour. Clinical stage was available or partially derivable for three patients: case 0006 was classified as Stage I (cT2N0M0), 0010 as Stage IVA (cT3N1bM1a), while 0002 had an IIA tumour and metastatic information.

**Table 1.** Demographic, clinical and histopathological characteristics of the five patients included in the colorectal cancer whole-genome sequencing pilot study. Patient identifiers (ID_g), Age range at consent (years), in 5-year intervals, biological sex, mismatch repair (MMR) status, family history of cancer, tumour site, clinical stage according to the American Joint Committee on Cancer (AJCC), 8th edition, histological grade and histological stage are presented for each participant. Tumour site is reported using the recorded anatomical description and ICD-10 code. Clinical stage was derived from radiological T, N and M classifications. ID 2 was classified as Stage IIA* based on the T3 category, assuming N0 and M0; however, the overall clinical stage could not be determined because radiological N and M classifications were unavailable. ID 6 was classified as Stage I (cT2N0M0), and ID 10 as Stage IVA (cT3N1bM1a). Clinical stage could not be assigned for ID 5 and ID 9 because TNM information was unavailable. Histological grade are reported as recorded; these entries reflect the original pathology descriptions and have not been reclassified. NOS : not otherwise specified.

| ID_g | Age range | Sex | MMR_group | Family_history | Tumour_site | Clinical_stage_AJCC8 | Histological.grade | Histological.stage |
|---|---|---|---|---|---|---|---|---|
| **0002** | 60-65 | Male | pMMR | Yes | C18.0 Caecum | Stage IIA* | Low Grade moderately differentiated adenocarcinoma NOS |  |
| **0005** | 35-40 | Female | Unknown | No | Splenic flexure tumour | NA | Invasive moderately differentiated adenocarcinoma |  |
| **0006** | 50-55 | Female | Unknown | Unknown | C18.0 Caecum | I | Invasive low-grade adenocarcinoma | Stage 1 |
| **0009** | 30-35 | Female | Unknown | No | Caecum tumour | NA | Invasive moderately differentiated adenocarcinoma |  |
| **0010** | 70-75 | Female | pMMR | No | C18.7 Sigmoid colon | IVA | Low Grade moderately differentiated adenocarcinoma NOS | low grade, moderately differentiated |

MMR status was available for two individuals, and both were classified as mismatch repair-proficient (pMMR), with retained expression of MLH1, PMS2, MSH2 and MSH6. One individual was recorded as not tested for MMR, while MMR information was missing for the remaining two. No MSI-PCR or *BRAF* V600E results were available: testing was recorded as not performed in three individuals, and information was missing for the other two.

### Quality analysis

Tumour DNA concentrations ranged from 76.4 to 159 ng/µL, corresponding to estimated total yields of 7.64–15.9 µg per sample, assuming a 100 µL elution volume. All samples met the laboratory’s DNA acceptance criteria (concentration ≥12 ng/µL and A260/280 ratio of 1.8–2.0). Estimated sequencing depth ranged from 49.2× to 82.4× for tumour samples and from 22.7× to 40.7× for matched normal samples. The median percentage of well-aligned reads across all samples was 96.3%. Following somatic variant calling and filtering, the number of retained somatic mutations ranged from 1,106 in sample 0005 to 65,379 in sample 0006 (table 2).

**Table 2.** Whole-genome sequencing metrics and somatic mutation burden across five paired colorectal tumour–normal samples. Sequencing yield (Gb) represents the total number of bases generated per sample, while sequencing depth (×) indicates the estimated mean genome coverage. The percentage of well-aligned reads represents the proportion of sequencing reads successfully aligned to the human reference genome (GRCh38). Somatic mutations after filtering correspond to the number of high-confidence somatic variants retained following variant calling and filtering using the CaVEMan and Strelka2 workflows. Tumour and matched normal samples are shown separately.

| Sample ID | Tumour yield (Gb) | Tumour depth (×) | Normal yield (Gb) | Normal depth (×) | Tumour well aligned (%) | Normal well aligned (%) | Somatic mutations after filtering |
|---|---|---|---|---|---|---|---|
| **0002** | 255.3 | 82.4 | 71.6 | 23.1 | 96.1 | 96.4 | 13612 |
| **0005** | 184.1 | 59.4 | 126 | 40.7 | 95 | 96.4 | 1106 |
| **0006** | 152.6 | 49.2 | 70.5 | 22.7 | 96.9 | 96.4 | 65379 |
| **0009** | 194.5 | 62.7 | 123.9 | 40 | 95.1 | 95 | 28901 |
| **0010** | 211 | 68.1 | 94 | 30.3 | 96.7 | 94.7 | 6580 |

### Somatic candidate mutations

Three of the five tumour samples (60%) harboured at least one somatic single-nucleotide variant (SNV) with a predicted protein-altering effect. Overall, 11 candidate variants were identified, comprising seven missense and four splice-site variants. These variants affected at least seven genes, including *COA7, EMB, NKPD1, POLR2A, SEPTIN3, and THSD1,* as well as three genes that could not be annotated (Figure 1).

**Figure 1.**
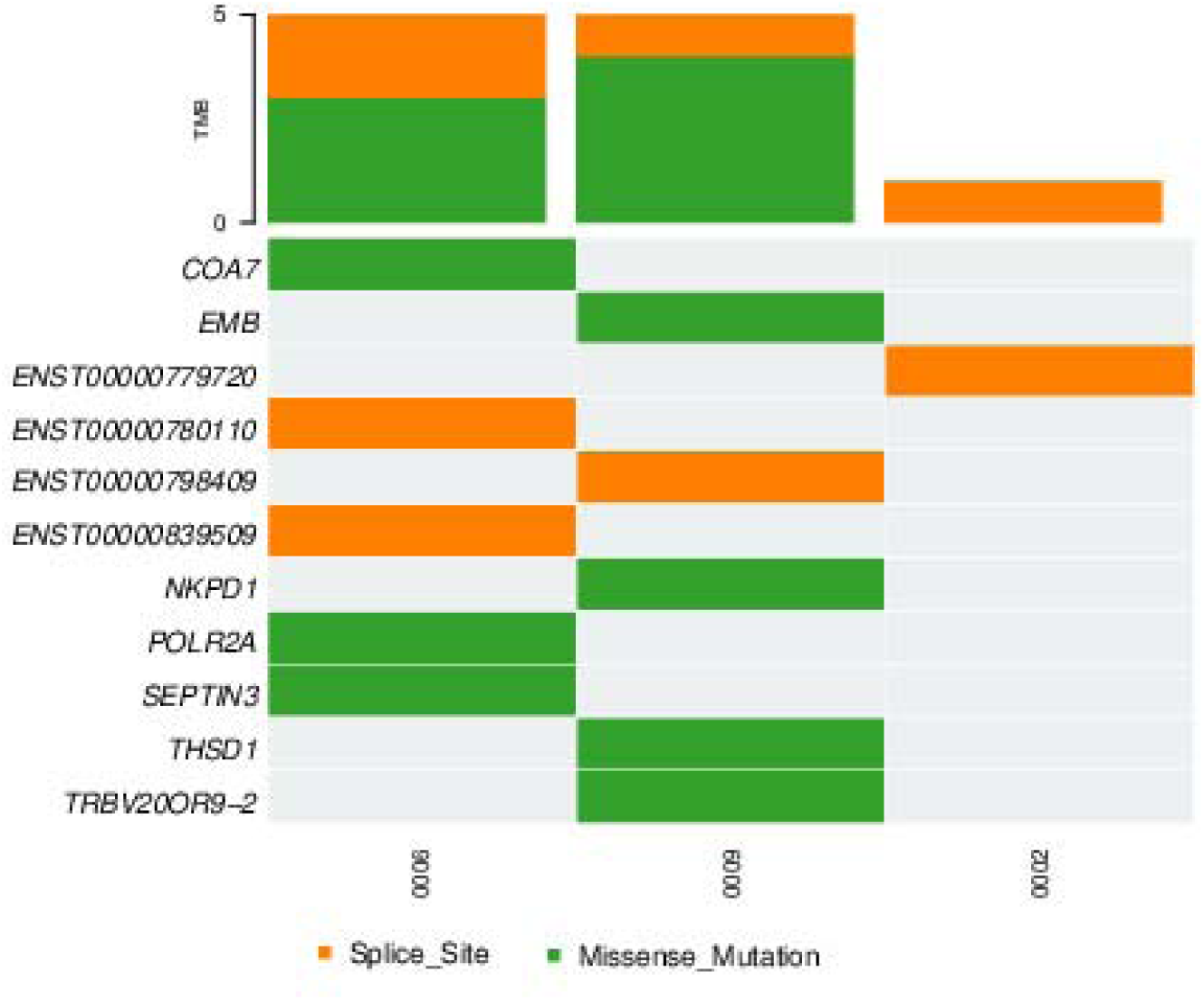
Somatic mutation profile of colorectal cancer tumours in the MOSAIC pilot study. Oncoplot showing the distribution of somatic protein-altering mutations across three tumour samples. Each row represents a mutated gene, and each column represents an individual tumour. Green indicates missense mutations, orange indicates splice-site mutations, and grey indicates no mutation detected in the corresponding gene and sample. The upper bar plot indicates the number of mutations per samplee. Overall, three tumour samples harboured candidate protein-altering variants affecting six genes (COA7, EMB, NKPD1, POLR2A, SEPTIN3 and THSD1), including seven missense and four splice-site variants.

**Figure 2.**
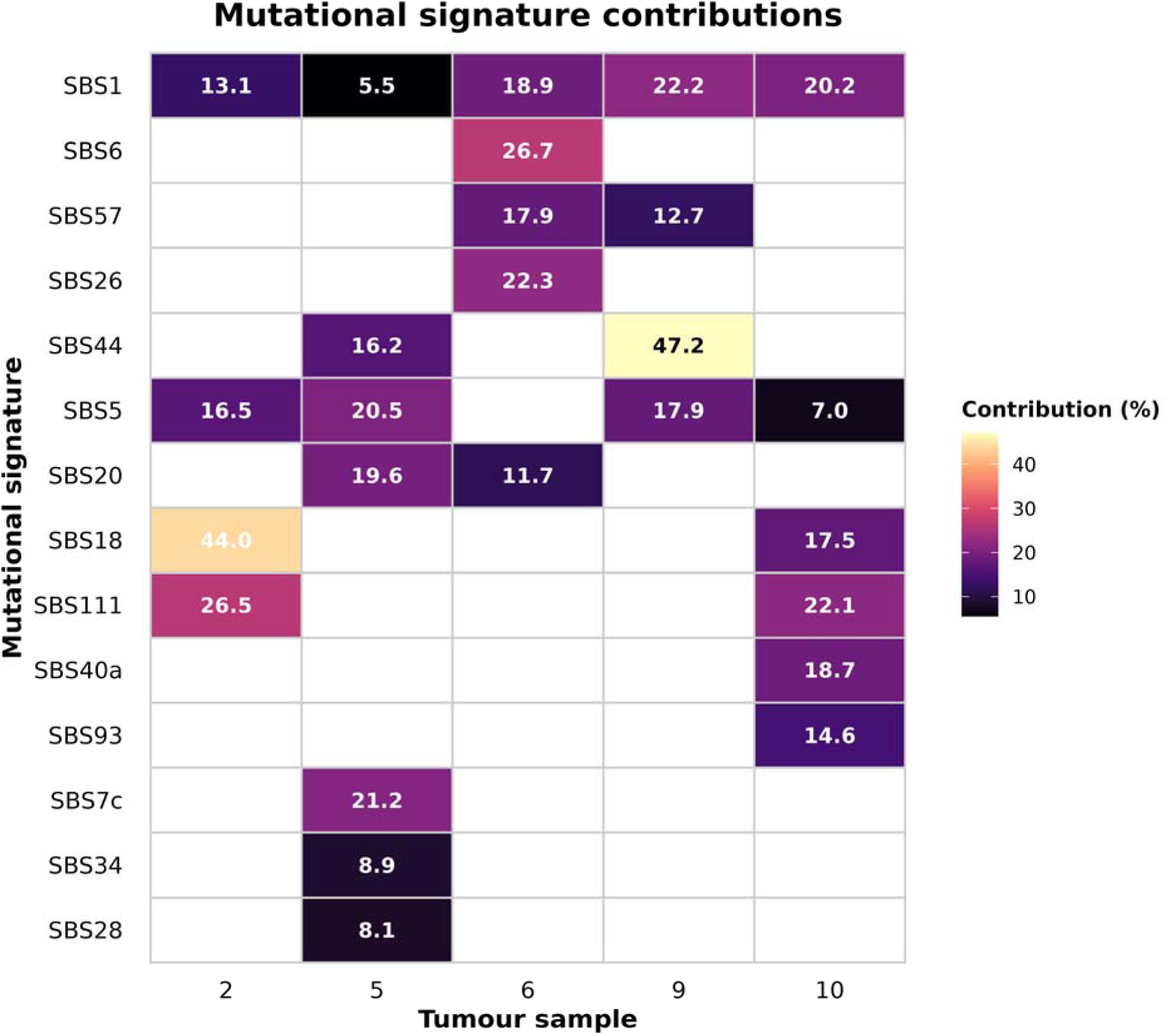
Mutational signature profiles across five colorectal cancer tumours from the MOSAIC pilot study. Heatmap showing the relative contribution (%) of candidate single-base substitution (SBS) mutational signatures identified in five colorectal cancer tumour samples (samples 2, 5, 6, 9 and 10). Each tile represents the percentage of mutations attributed to an individual mutational signature within a tumour. Colour intensity indicates the relative contribution of each signature, with higher contributions represented by brighter colours. White tiles indicate signatures that were not retained in the candidate signature table (contribution below the 5% selection threshold or not detected). Percentages are displayed within tiles.

### Mutational signature

Mutational signature analysis revealed considerable heterogeneity in the mutational profiles across the five colorectal cancer tumours. SBS1 was identified in all five samples and SBS5 in four representing shared age-related and clock-like mutational processes, respectively.

Additional signatures varied between tumours. SBS18, associated with oxidative damage, contributed substantially to the mutational profile of sample 0002 (43.96%) and was also detected in sample 10 (17.48%). SBS44, associated with defective DNA mismatch repair (MMR), was identified in samples 0005 and 0009, accounting for 16.18% and 47.16% of their respective mutational burdens. Sample 0006 exhibited a distinct profile characterised by the presence of SBS6 (26.73%), SBS26 (22.33%) and SBS20 (11.74%), all associated with deficient MMR-related mutational processes associated with mutation burden higher (table 2). SBS20 was additionally identified in sample 0005 (19.62%). SBS57 makes up 13–18% of the profile in samples 0006 and 0009 but it is likely an artefact see https://cancer.sanger.ac.uk/signatures/sbs/sbs57/.

Overall, these findings suggest inter-individual heterogeneity in the mutational processes contributing to colorectal cancer development, with several tumours exhibiting signatures associated with defective DNA mismatch repair. The identification of these signatures warrants further investigation alongside molecular characterisation of MMR status (Figure 3).

## Discussion

This pilot study represents a collaborative effort involving the University of the Witwatersrand, CHBA, SBIMB, the Wits DIH, WDGMC, the University of Bristol and the IARC. Through this collaboration, tumour and matched peripheral blood samples were collected from Black South African patients with CRC, followed by WGS, sequencing quality control and preliminary somatic variant and mutational signature analyses. This work provides an initial characterisation of somatic genomic alterations in a population that remains underrepresented in large-scale CRC genomic studies.

A total of five individuals were phenotyped and successfully included in the sequencing analysis, with clinical, pathological and demographic information collected through the CoRect Cancer study. Molecular characterisation included the assessment of mismatch repair (MMR) status where immunohistochemistry results were available. Additional MSI and MMR testing was not routinely performed on biopsy specimens within South African state pathology services and would therefore have required dedicated testing within the study. Although whole-genome sequencing provides opportunities for the computational assessment of MSI and MMR-associated mutational processes, complementary pathological evaluation remains important for molecular characterisation. Future studies will benefit from dedicated resources to support additional pathological and molecular testing, enabling more comprehensive integration of genomic, clinical and epidemiological data.

Three of the five tumour samples harboured at least one candidate somatic variant with a predicted protein-altering consequence. Across these samples, seven missense and four splice-site variants were identified in eleven genes. These variants represent potential functional candidates; however, their contribution to CRC development and progression remains to be established. Of particular interest, POLR2A is located adjacent to the tumour suppressor gene TP53 on chromosome 17p13 and has previously been reported to be co-deleted with TP53 in CRC, potentially creating a therapeutic vulnerability (Liu et al., 2015). Nevertheless, the identification of a protein-altering variant in POLR2A does not establish a functional or oncogenic role, and further investigation would be required to determine its biological significance.

Interestingly, the two samples without identified protein-altering candidate variants also exhibited comparatively lower total numbers of detected somatic mutations. This observation could reflect differences in tumour biology and mutational burden, potentially influenced by tumour stage or mismatch repair status. Alternatively, differences in tumour purity, intratumoural heterogeneity and sequencing sensitivity may have contributed to the lower number of detected variants. Further studies incorporating tumour purity estimates, sequencing sensitivity assessments and comprehensive clinical annotation will be necessary to investigate these observations.

Mutational signature analysis identified several processes previously reported in CRC. These included age-associated or clock-like signatures SBS1 and SBS5, which are known to accumulate with age, as well as SBS18, associated with oxidative damage. Other identified signatures included SBS20, SBS26 and SBS44, which have been reported in colorectal cancer in association with particular DNA repair and mismatch repair-related processes and suggest that patient 0006 is MMR deficient with a highest burden mutation suggest a MSI but that should be confirmed by further test . Notably, SBS111, a recently characterized signature provisionally associated with DNA repair infidelity in CRC, was also identified in our analysis. Its recent description in microsatellite-stable CRC highlights the importance of considering emerging mutational processes when interpreting somatic mutation profiles (Institute, 2026; Kazachkova et al., 2026; Liu et al., 2015). However, the presence of these signatures should not be interpreted as definitive evidence of a specific underlying DNA repair defect, particularly in the absence of MSI/MMR characterisation and confirmatory molecular analyses. We did not identify SBS88, a mutational signature associated with exposure to colibactin, a genotoxic compound produced by certain *Escherichia coli* strains carrying the pks pathogenicity island. Colibactin-associated mutagenesis has attracted increasing attention in CRC research, particularly because of its association with early-onset disease. Recent large-scale investigations have demonstrated geographical and age-related variation in mutational processes, including the prevalence of colibactin-associated signatures in CRC (Díaz-Gay et al., 2025). Although SBS88 has been reported in a proportion of CRC cases, its prevalence varies across populations and study cohorts. Its absence in our five samples should therefore be interpreted cautiously, as the limited sample size and sensitivity of signature detection may have prevented its identification. Nevertheless, the contribution of colibactin-associated mutagenesis to CRC in sub-Saharan Africa, and particularly Southern Africa, remains an important question that warrants investigation in larger population-based cohorts.

Finally, the age range of our participants (34–73 years) represents another important consideration when interpreting the observed mutational profiles. Age is an important determinant of somatic mutation accumulation, with signatures such as SBS1 and SBS5 showing age-related patterns. Furthermore, molecular and clinicopathological characteristics may differ between early-onset and later-onset CRC, although these differences are not necessarily explained by age alone (Kazachkova et al., 2026). Recent genomic investigations have further highlighted the contribution of age and geographical variation to the mutational landscape of CRC (Storandt et al., 2025). Nevertheless, these findings reinforce the importance of investigating age-related and population-specific mutational processes in African CRC populations.

### Limitations and future directions

This study represents an initial step towards characterising the somatic genomic landscape of CRC in Black South African patients. Several limitations should be acknowledged. The pilot cohort comprised only five individuals, limiting statistical power. The current analysis primarily focused on single-nucleotide variants (SNVs) and their predicted functional consequences, and the systematic assessment of germline susceptibility has not yet been performed.

The findings should be considered exploratory rather than representative of the broader South African CRC population. Larger cohorts with comprehensive pathological, clinical and molecular characterisation will be essential to validate the candidate variants, investigate mutational signature distributions and better understand the contribution of population-specific genetic, environmental and microbial exposures to CRC development in sub-Saharan Africa.

An objective of the MOSAIC collaboration is to build on this pilot by expanding recruitment across Africa and increasing sample sizes to enable a more comprehensive investigation of the molecular landscape and aetiology of CRC. These efforts aim to better understand the molecular drivers of CRC across African populations, including differences between early-onset and later-onset disease, and ultimately inform population-specific risk assessment and precision oncology research.

## Ethics approval and consent to participate

Ethical approval for the study was obtained from the University of the Witwatersrand, Johannesburg, South Africa (M250822, M240947, M2409115, BEC20240904), as well each respective clinical site. Written informed consent was obtained from all participants prior to sample collection. Consent included permission for the collection, storage, and future use of biological samples and associated data for health-related research, subject to approved ethical and governance requirements.

## Data Availability

The data supporting the findings of this study are available from the corresponding author (JTB) upon reasonable request.

## Authors’ contributions

JTB, PCH and PS conceived and designed the study. RMM and PCH secured funding. BB, NH, VS, JF and LC contributed to sample identification, recruitment and data collection. ED and SW performed the sequencing. JTB conducted the bioinformatics analyses and drafted the manuscript. All authors contributed to the interpretation of the results, critically reviewed the manuscript, and approved the final version.

## Members of the Molecular, Somatic and Infectious Drivers of Colorectal Cancer in Africa (MOSAIC) collaboration

### Byline authors

Jean-Tristan Brandenburg, Lisa Campbell, Dewaldt Engelbrecht, Christopher G. Mathew, Richard M Martin, Brendan Bebington, Vuyo Soldati, Sandra Perdomo, Wendy Stevens, June Fabian, Philip C Haycock

### Non-author collaborators

Michele Ramsay, Wenlong Carl Chen, Moses Koobotse, Matthew Suderman, Paul D Yousefi , Thomas Vogl, Mandeep Kaur, Adam Chambers, Helen Winter, Boitumelo Ramasodi, Abel David González-Pérez, Nadine Harran, Jennifer Groverman, Nicole Sheahan, Dr Miguel Vanterpool Héctor, Lisa Ware, Olusegun Isaac Alatise

## Acknowledgements

We thank all study participants who contributed to the respective datasets. We acknowledge the contributions of Prof Scott Hazelhurst for access to the Wits computer cluster, the SBIMB (Sydney Brenner Institute for Molecular Bioscience) biobank and personnel managing the samples, the contributions of our research assistants, , laboratory scientists, administrators, data personnel, and all other staff who contributed to the data and sample collections, processing, and storage.

## Disclaimer

Where authors are identified as personnel of the International Agency for Research on Cancer/World Health Organization, the authors alone are responsible for the views expressed in this article and they do not necessarily represent the decisions, policy or views of the International Agency for Research on Cancer/World Health Organization.

